# Estimating the Risk-Based Value of the Diabetes Prevention Program: How well does clinical trial-based cost-effectiveness apply to the real world?

**DOI:** 10.1101/2025.10.02.25337195

**Authors:** Natalia Olchanski, Samuel B. Weidner, Ching-Hsuan Lin, David M. Kent

**Affiliations:** Center for the Evaluation of Value and Risk in Health, Institute for Clinical Research and Health Policy Studies, Tufts Medical Center, 800 Washington St, #063, Boston, MA 02111; Department of Medicine, Tufts University School of Medicine, 145 Harrison Ave, Boston, MA 02111; Flatiron Health, Data Insights, New York, NY; Institute for Clinical Research and Health Policy Studies, Tufts Medical Center, 800 Washington Street #63, Boston, MA 02111, USA

## Abstract

**Objectives:** Many economic evaluations rely on clinical trial data that may not represent real world populations and intervention effectiveness. We compare risk and cost-effectiveness for the original Diabetes Prevention Program (DPP) clinical trial population and a real world population eligible for the National Diabetes Prevention Program (NDPP).

**Methods:** We identified National Health and Nutrition Examination Survey (NHANES) subjects eligible for the NDPP and adjusted projections using survey weights to produce real world (US population) representative results. We used clinical predictive models to estimate individual diabetes risk and microsimulation to estimate lifetime costs, benefits, and net monetary benefits (NMB) for lifestyle intervention and metformin. We compared results across the original DPP clinical trial and real world populations.

**Results:** Only 20% of the NHANES population eligible for NDPP met inclusion/exclusion criteria for the DPP trial. Three-year risk of diabetes onset for trial population (mean 19.7%, median 10.3%) exceeded corresponding risk for the NHANES population (mean 14.6%, median 4.8%). The proportion of individuals with < 10% three-year diabetes risk for the trial population (49%) was less than the corresponding proportion for NHANES (67%). Lifestyle intervention had mean NMB $34,889 for the DPP trial population and $28,652 for NHANES.

**Conclusions:** Real world populations eligible for the NDPP include a greater proportion of low-risk individuals, for whom prevention programs may confer smaller benefits. Using individualized diabetes risk estimates to inform referrals and prioritization for diabetes prevention can maximize benefit and is expected to have greater impact on real world populations than the clinical trial cohort.

**Key Points:** Real world populations eligible for the National Diabetes Prevention Program include a greater proportion of low-risk individuals than the original clinical trial, and for these people, prevention programs may confer smaller benefits. Using individualized diabetes risk estimates to inform referrals and prioritization for diabetes prevention can maximize benefit and is expected to have greater impact on real world populations than the clinical trial cohort.

## 1. Introduction

More than 1 in 3, or approximately 98 million American adults have prediabetes [1], putting them at an increased risk of developing type 2 diabetes. Among persons with prediabetes, an individual’s risk of developing diabetes depends on many factors and has been shown to vary widely [2]. While generally considered effective, the National Diabetes Prevention Program (NDPP) has struggled to reach high risk populations and engage individuals in a widespread uptake of the program [3]. Due to the large size of the US eligible population, effective and equitable outreach and engagement strategies require substantial resources. While it is not feasible to enroll everyone with prediabetes in the DPP, knowing individualized risks of diabetes and the potential benefits of preventive interventions might help health care providers to prioritize patients for preventative interventions and to optimize the number of type 2 diabetes cases prevented [4–6].

Many economic evaluations rely on clinical trial data, but these data may not be representative of real world clinical practice and outcomes. Clinical trials often include populations that are different from those who receive the interventions after market approval and dissemination. For example, NDPP has different inclusion/exclusion criteria than the original clinical trial of the DPP intervention [7]. In the case of prediabetes, the effectiveness and value of the intervention is highly dependent on the individual’s characteristics and the risk of developing type 2 diabetes [6, 8].

Because prediabetes is a broad category with people at vastly different risk levels, it is not clear how the clinical trial population compares with the real world population receiving DPP, and whether economic analysis based on clinical trial data is useful for evaluating a real world intervention like NDPP. In this study we compare the original DPP clinical trial population to US individuals eligible for NDPP and the economic value of DPP between these populations. We also examine how the value of using individualized risk prediction for prioritizing enrollment into DPP translates between clinical trial and real world populations.

## 2. Methods

We compared the demographic characteristics, type 2 diabetes risk factors, and the US Centers for Disease Control and Prevention (CDC) prediabetes risk scores between a clinical trial population and a real world population of individuals with prediabetes who fit eligibility criteria for NDPP. We grouped the populations into five quintiles by three-year type 2 diabetes risk. Our analysis utilized the “net monetary benefit” (NMB) metric to compare the economic value of two DPP strategies, including intensive lifestyle intervention and metformin. Net monetary benefit is the difference between the monetized value of health gains associated with an intervention and the net healthcare costs associated with the intervention. We estimated the lifetime health benefits and costs of the strategies using simulation modeling. We then defined the monetized value of health as the product of quality-adjusted life expectancy gain associated with an intervention and the value of quality-adjusted life years (QALYs), which we set as $100,000/QALY in our base case [9].

Our second analysis examined the value of using individualized diabetes risk information to prioritize enrollment in a diabetes prevention program. To do this, we simulated and compared the NMB of preventative interventions for two scenarios: (1) treating the 20% of the population at the highest risk of developing diabetes, and (2) treating a random 20% sample of the population eligible for National DPP. We projected these scenarios in both the real world population and the clinical trial cohort. Next, we compared the mean NMB per treated patient between the real world population and the clinical trial population. Statistical analyses were performed using STATA version 17 and R 4.2.0. (R Core Team, 2022)

### 2.1 Data and Population

Our clinical trial population was based on the original Diabetes Prevention Program clinical trial, a randomized, controlled clinical trial conducted at 27 sites around the United States from 1996-2001 [7]. The trial enrolled individuals older than 25 who had impaired glucose tolerance, as well as a BMI ≥ 24. Impaired glucose tolerance was defined as having both a fasting plasma glucose (FPG) test within the range of 95-125 mg/dL and a two-hour oral glucose tolerance test (OGTT) within the range of 140-199 mg/dL.

We established our *real world population* of individuals with prediabetes who fit eligibility criteria for NDPP using data from the CDC’s National Health and Nutrition Examination Survey (NHANES), a national survey conducted every two years that combines interviews and physical/lab examinations to “assess the health and nutritional status of adults and children in the United States” [10]. Taking data from two NHANES waves (the 2015-16 wave, and the 2017-18 wave), we applied the eligibility criteria from the NDPP to select our population (Table 1). We used NHANES survey weights to weight the NHANES study cohort to represent a national population eligible for DPP (95,469,375 Americans). Additionally, we assessed our NHANES cohort of individuals eligible for NDPP to estimate the proportion who would have been eligible for inclusion in the original DPP clinical trial.

**Table 1:** Side by side comparison of selected inclusion/exclusion criteria for the Diabetes Prevention Program (DPP) clinical trial, the National DPP program, and the cohort for derivation of the Tufts PACE diabetes risk prediction model

| Variable | Inclusion Criteria |  |  |
| --- | --- | --- | --- |
|  | *Diabetes Prevention Program NIDDK trial* | *National Diabetes Prevention Program* | *Tufts PACE DPP risk model derivation population* |
| Age | 25+ | 18+ | 25-75 (upon index visit) |
| BMI | >=24; >= 22 if Asian | >= 25; >=23 if Asian | N/A |
| Hemoglobin A1c | N/A | 5.7-6.4% inclusive | 5.7-6.4% inclusive |
| Fasting plasma glucose | 95-125 mg/dL inclusive | 100-125 mg/dL inclusive | 100-125 mg/dL inclusive |
| 2-hour glucose tolerance test | 140-199 mg/dL | 140-199 mg/dL inclusive | N/A |
| Gestational diabetes | N/A | Have a previous diagnosis of gestational diabetes | N/A |
| CDC prediabetes risk test | N/A | Receive a screening result of high risk for type 2 diabetes | N/A |
| Exclusion criteria | List of 39 exclusions as to not influence the study | Pregnancy | Pregnancy, > 200 mg/dL random glucose, lack of clinical activity |

**Note:** CDC, Centers for Disease Control and Prevention

The current NDPP developed by the CDC, based on the results of the original DPP trial, has similar inclusion criteria as the trial with some differences (Table 1). To be eligible for the NDPP, you must be at least 18 years of age, have a BMI ≥ 25 (23 for Asians), and have at least one of the following: a hemoglobin A1c (HbA1c) blood test within the range of 5.7%-6.4%, a FPG test within the range of 100-125 mg/dL, an OGTT within the range of 140-199 mg/dL, a previous diagnosis of gestational diabetes, or a screening result of high risk using the CDC Prediabetes Risk Test tool (i.e., a score of 5 or higher [11]).

### 2.2 Estimating Costs and Benefits

We used a previously constructed decision-analytic microsimulation model [6] to project costs of care and health outcomes for individuals with prediabetes eligible for the NDPP over a lifetime time horizon (Fig.1). Our model calculated lifetime costs and QALYs for each individual as they progressed from “Prediabetes” health state to “Type 2 Diabetes” or “Death” health states. The risk model estimated individual diabetes progression probabilities based on baseline characteristics. For those who progressed to “Type 2 Diabetes”, we estimated the trajectories of costs and outcomes after the onset of diabetes using the Michigan Model for Diabetes (MMD) [12, 13], available at https://diabetes.med.umich.edu/michigan-model-diabetes. For each individual, the model predicted costs from the healthcare sector perspective and QALYs for two diabetes prevention strategies, lifestyle intervention used in NDPP and metformin, as well as for usual care. We used previously described model parameter inputs for costs, health state utilities, and event and disease progression rates after diabetes onset [6].

**Figure 1:**
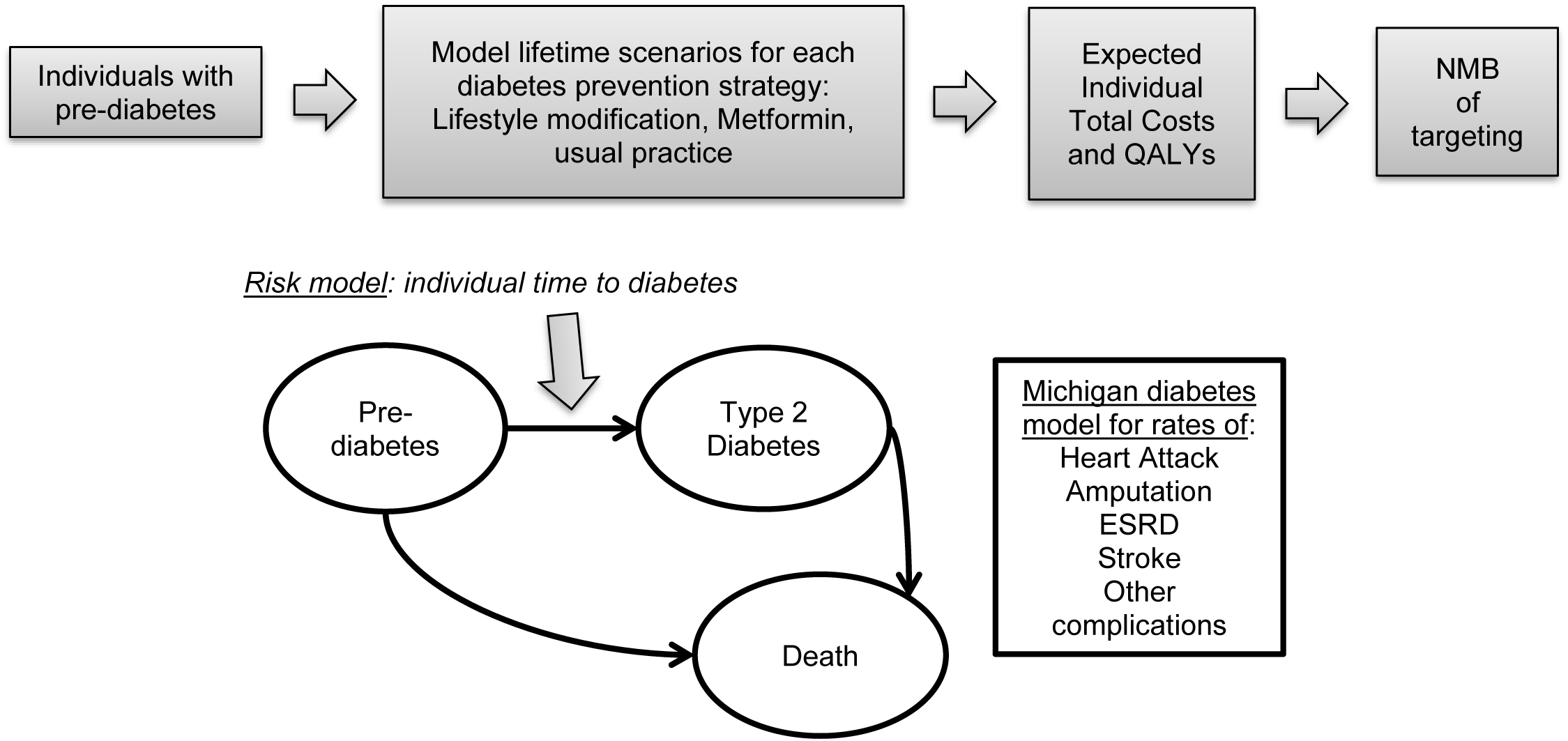
Simulation model overview. Adapted from Olchanski N, van Klaveren D, Cohen JT, Wong JB, Ruthazer R, Kent DM. Targeting of the diabetes prevention program leads to substantial benefits when capacity is constrained. Acta Diabetol. 2021;58(6):707-22.

We applied the model to the Diabetes Prevention Program (DPP) Clinical Trial population (n = 3068) and to the real world population drawn from NHANES (n = 4387). Using NHANES survey weights we weighted the NMB, costs, and QALYs for the NHANES-based cohort to represent a national population eligible for NDPP (95,469,375 Americans). We discounted costs and QALYs using a 3% discount rate and adjusted all costs to 2020 dollars using the Personal Consumption Expenditure Health Index (PCE – Health) and the Personal Health Care Index (PHC) [14]. We used R (R Core Team, 2020) and RStudio (Rstudio Team, 2020) for modeling and statistical analysis.

### 2.3 Risk and Progression to Type 2 Diabetes

To predict individual risk of developing diabetes over time under each treatment strategy for both our study populations of adults with prediabetes, we utilized a Cox proportional hazards model. The model used as inputs baseline characteristics of each individual, along with the specified treatment strategy of lifestyle intervention, metformin, or usual care.

Our Cox proportional hazards model used the Tufts Predictive Analytics and Comparative Effectiveness DPP Risk model (Tufts PACE DPP risk model) to predict 3-year risk of diabetes. The Tufts PACE DPP risk model was a previously published diabetes onset prediction model developed and validated using electronic health record (EHR) data from the Optum Labs Data Warehouse [15]. Briefly, it used eleven variables expected to be reliably obtained from EHR records (age, gender, race, smoking-status, BMI, presence or absence of a hypertension diagnosis, systolic blood pressure, HDL cholesterol, triglycerides, fasting plasma glucose, and HbA1c). The model predicted individual 3-year risk of diabetes onset using baseline characteristics, while also estimating treatment effects of lifestyle intervention and metformin at different diabetes risk levels using data from the DPP clinical trial.

We extended our Cox proportional hazards model beyond 3 years using long term patient outcome data collected in the DPP Outcomes Study (DPPOS) [16], an open label follow-up study conducted over seven years for a subset of the participants in the DPP clinical trial. Our Cox proportional hazards model included the linear predictor from the Tufts PACE DPP risk model and indicator variables for the treatment arm to predict long term diabetes risk in the lifestyle intervention arm. We then estimated the placebo and metformin risk trajectories by applying treatment effect hazard ratios to the lifestyle arm. The base case made a conservative assumption of no additional treatment effect of lifestyle intervention or metformin after the initial 3 years.

In order to apply the risk prediction model to the real world population, we imputed missing prediction factors in two cases. First, NHANES procedure randomly assigns participants to either morning or afternoon sessions, and only those in the morning sessions give a fasting blood sample [17] to measure fasting plasma glucose and triglycerides. Because the data are missing at random due to the NHANES random assignment, we used single imputation to impute the missing values for fasting plasma glucose and triglycerides. The second use of imputation was for variables where rates of missingness were less than 5%, including HbA1c, systolic blood pressure (SBP), and HDL cholesterol.

## 3. Results

The two study populations had similar age distributions, but the clinical trial cohort included a higher proportion of women and a higher proportion of Black individuals than the modeled real world US population eligible for National DPP based on NHANES (Table 2).

**Table 2:** Study population characteristics.

|  | **DPP Clinical Trial population (N=3068)** | **US population eligible for National DPP program (NHANES n=4387, weighted N = 95,469,375)** |
| --- | --- | --- |
| **Demographic characteristics** |  |  |
| Age | 50.6 (52.0) | 52.5 (54.0) |
| Female | 66.7% | 46.4% |
| Race |  |  |
| Non-Hispanic White | 57% | 63.2% |
| Non-Hispanic Black | 21% | 11.7% |
| Hispanic | 16% | 15.8% |
| Asian | N/A | 4.7% |
| Other or Missing | 5% | 4.7% |
| **Clinical/health behavior history** |  |  |
| Body Mass Index (BMI) | 33.5 (33) | 32.7 (31.2) |
| Systolic blood pressure | 124.1 (122) | 127.4 (125.3) |
| Hypertension (yes/no) | 27% | 41.8% |
| Smoking status |  |  |
| Never | 57.1% | 54.0% |
| Former | 33.8% | 29.1% |
| Current | 9.1% | 16.8% |
| Previous diagnosis of gestational diabetes (yes/no) | 10.6% | 1.5% |
| Physical activity (yes/no)* | 27.8% | 60.6% |
| CDC prediabetes risk calculator score | 5.5 (5.0) | 5.5 (5.0) |
| Proportion above high risk cutoff using CDC prediabetes risk predictor | 73.5% | 77% |
| **Lab values** |  |  |
| Hemoglobin A1c | 5.9 (5.9) | 5.6 (5.6) |
| Fasting plasma glucose (FPG) | 107.2 (105) | 104.9 (105.0) |
| Oral glucose tolerance test (OGTT) | 164.7 (163) | 119.6 (116.0) |
| High-density lipoprotein (HDL) cholesterol | 45.6 (44) | 51.8 (49.0) |
| Triglycerides | 162.9 (141) | 123.4 (102.0) |

Notes: Data are presented as mean (median) for continuous measures, and % for categorical measures. CDC, Centers for Disease Control and Prevention; DPP, diabetes prevention program; NHANES, National Health and Nutrition Examination Survey.
\* Physical activity shows proportion of physically active individuals defined as follows:
- NHANES population: coded as physically active if they had 150 or more minutes per week of moderate or vigorous physical activity[27]. - DPP Clinical Trial population: described as more physically active when compared with others at the same age.

We found that the clinical trial population had a higher diabetes risk profile than the real world population at every risk quintile (Fig. 2). Only 20% of the real world population met inclusion/exclusion criteria of the original DPP clinical trial. Using the Tufts PACE DPP diabetes risk prediction model, we found the mean three year risk of diabetes onset for the clinical trial population to be 19.7% with a median of 10.3%, while the real world population had a mean of 14.6% and a median of 4.8%. Individuals with three year diabetes risk under 10% made up 33% of the clinical trial cohort and 66% of the real world population.

**Figure 2:**
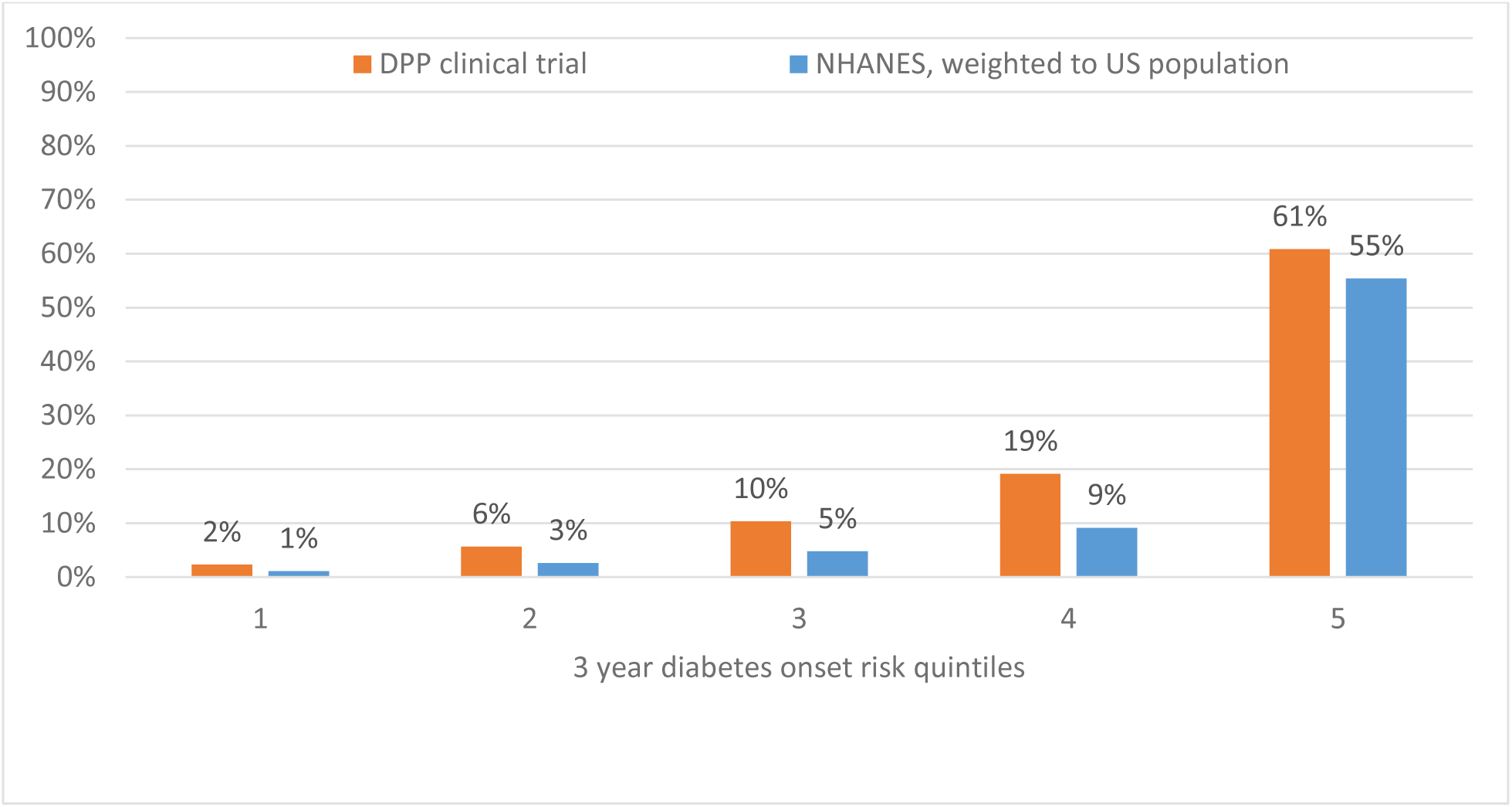
Comparison of DPP clinical trial cohort and real world population eligible for national DPP (extracted from NHANES waves 2015-2018), 3 year risk of type 2 diabetes onset by risk quintile.

### 3.1 Value of Lifestyle intervention

Mean NMB of lifestyle intervention compared to usual care for the real world population ($28,652) was lower than the corresponding value for the clinical trial population ($34,889). NMB was positive in both study populations across diabetes risk quintiles. The mean NMB values for each diabetes risk quintile were lower for the real world population compared to the clinical trial population (Fig. 3).

**Figure 3:**
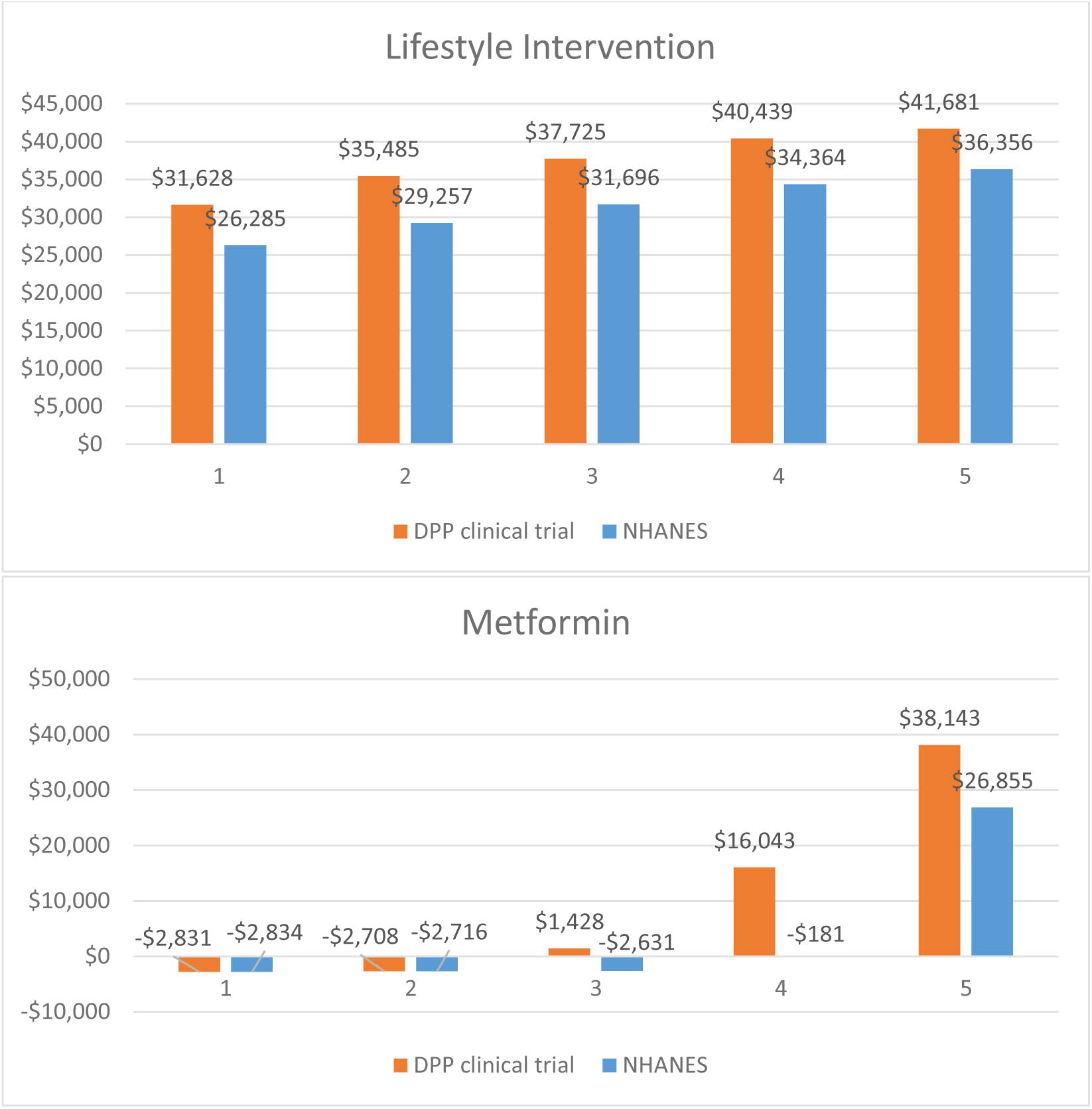
Net Monetary benefit of Metformin and Lifestyle intervention by diabetes risk quintile –comparison of DPP clinical trial cohort and real world population.

### 3.2 Value of Metformin intervention

Mean NMB for metformin compared to usual care for the real world population ($5,391) was lower than the corresponding value for the clinical trial population ($9,749). Mean NMB were lower for the real world population compared to the clinical trial population across all diabetes risk quintiles (Fig. 3). The proportion of people with NMB<0 was 67% for real world population and 49% for the clinical trial population, reflecting the proportions of low risk individuals in these populations.

### 3.3 Targeting diabetes prevention

In the case of total DPP program capacity of 20% of the eligible population (approximately 17 million people), targeting treatment to individuals in the highest diabetes risk quintile, which had the highest predicted NMB, increased average NMB per person (Fig. 4). The increases in NMB values were more substantial in the real world population compared to the clinical trial population (Fig. 4). For lifestyle intervention, the gain in average NMB was $7,895 (41%) per person in the real world population, compared to $5,753 (27%) per person in the clinical trial population. For metformin, the gain in average NMB was $11,152 (150%) per person in the real world population, compared to $10,631 (122%) per person in the clinical trial population (Fig. 4).

**Figure 4:**
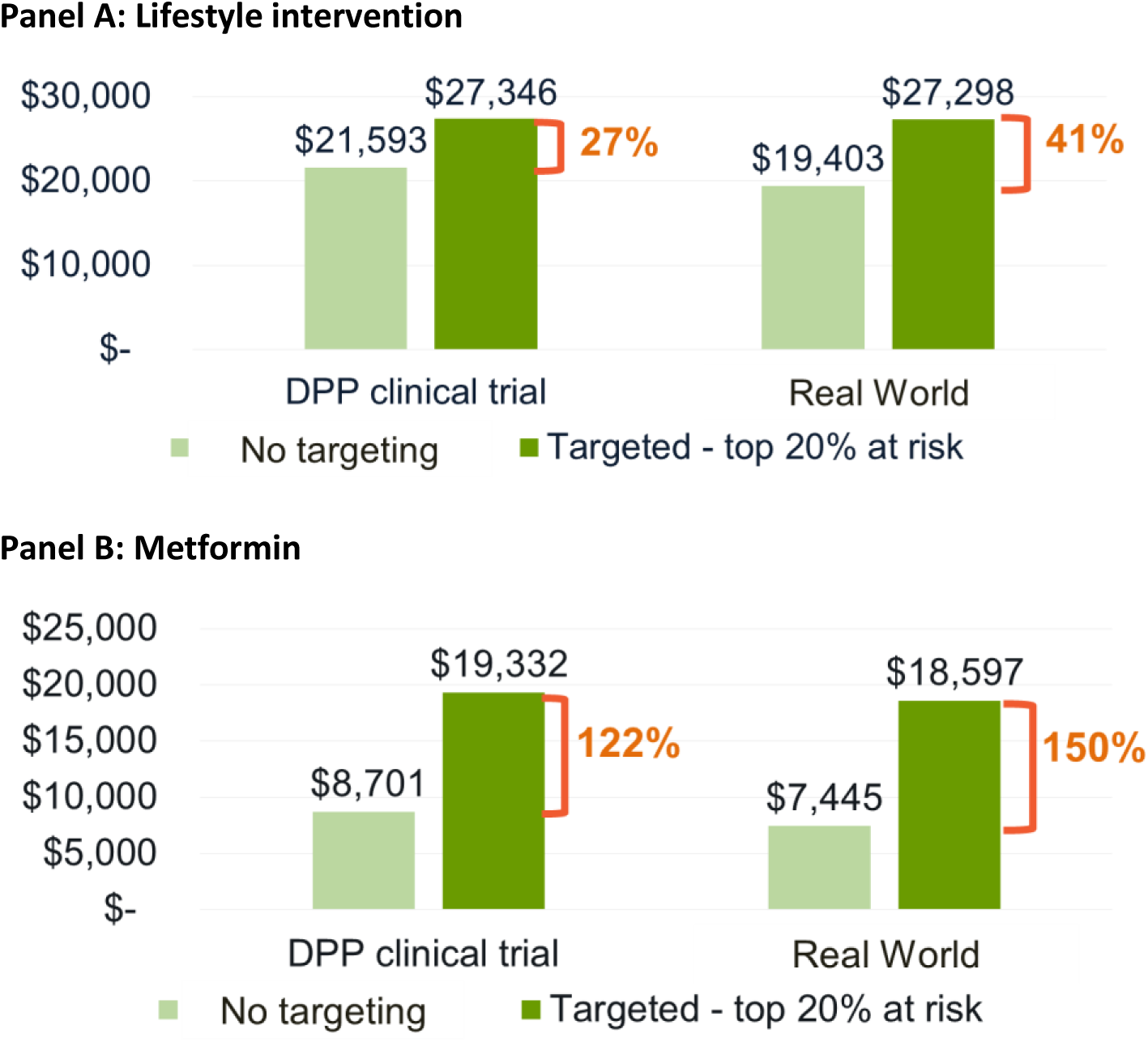
Value (Net Monetary Benefit) of prioritizing by diabetes onset risk – comparison of DPP clinical trial cohort and real world population.

## 4. Discussion

The focus of the current analysis was to evaluate the impact of using real world population data in an economic analysis of DPP, compared to a clinical trial cohort based assessment, which is typical of health economic evaluations. We found that real world population eligible for the NDPP included a greater proportion individuals with lower risk of diabetes onset, partly due to the differences in the less stringent eligibility criteria used by National DPP compared with the original DPP clinical trial. Additionally, there were demographic differences between these groups. These findings are consistent with a body of literature showing that clinical trials are not representative of real world treated populations [18]. Of the population we found in NHANES that is eligible for the National DPP, only 20.7% would have been eligible to participate in the original DPP clinical trial.

Our findings indicate that the real world NDPP eligibility criteria include lower risk individuals who benefit less from the lifestyle or metformin interventions. We found that for individuals with prediabetes in the lower risk quintiles, prevention programs conferred smaller benefits. Further we found that DPP conferred lower NMB in the real world population compared to the participants in the original DPP clinical trial. Consistent with our findings, Shahraz et al also found that the expansion of glycemic criteria for prediabetes since the original DPP clinical trial has reduced overall benefit and efficiency of the program [19]. Using the expanded criteria, the prevalence of prediabetes increased to 51.3% in the US [2]. Treating all individuals meeting these criteria with the current resources poses significant challenges.

Using individualized diabetes risk estimates to inform referrals and prioritization for diabetes prevention may have a higher impact among real world populations than based on the clinical trial cohort. We built our work on a prior study that demonstrated the benefits of prioritizing treatment based on diabetes risk [6]. Unsurprisingly, our model indicated that outcomes improved when interventions targeted individuals at the highest risk. However, the advantage of focusing on highest-risk populations was even more pronounced in real-world populations. This may be due to the clinical trial population being specifically selected for its likelihood to benefit, reducing the need for additional prioritization. In contrast, targeting higher-risk individuals for participation in the DPP is particularly sensible in resource-constrained real-world contexts [20]. In this light, using an EHR-compatible risk tool shows promise, as our risk scores were derived from data available in the clinical practice setting. While previous studies have demonstrated the benefits of EHR-based tools for diabetes management in inpatient settings [21], their impact in primary care is emerging [22]. Future research exploring the application of personalized diabetes risk and outcome prediction in community settings could have significant policy implications.

Our study has several limitations. First, we did not examine the impact of health care disparities. Disparities in diabetes care and outcomes by race and low income groups are well documented [23, 24]. Efforts to prioritize and improve the reach and uptake of DPP should consider and address the equity of access and resources to engage participants. Future economic evaluation should incorporate the impacts on health inequality, such as a distributional cost-effectiveness analysis [25]. Second, for this illustrative case study we performed an assessment from the health sector perspective, rather than including all possible impacts from the societal perspective, such as work productivity. Including broader elements of value [26] would have resulted in higher value of DPP across all analyses, especially in high risk individuals, and would amplify the value of prioritization by diabetes risk.

## 5. Conclusion

The real-world populations eligible for the NDPP had a greater proportion of low-risk individuals compared to those enrolled in the clinical trials of DPP. For these lower-risk individuals, the benefits of the prevention programs was smaller on average compared to those with higher risk. As a consequence, adopting a more tailored approach that leveraged individualized diabetes risk estimates to prioritize intervention had a greater impact on diabetes prevention efforts in real-world populations.

Our findings suggest that current technology assessments, which predominantly rely on clinical trial data, may overestimate the potential benefits of the NDPP when generalized to broader, more heterogeneous populations. Integrating data from real-world populations could ensure more accurate predictions of program effectiveness, optimize resource allocation, and ultimately enhance the overall impact of diabetes prevention strategies on public health. Technology assessments based on clinical trial data should be revised using real world population and treatment effect data. In addition, clinical trials can improve their applicability and relevance to real world practice by including more diverse participants and reporting risk-stratified results, enabling analysis of heterogeneity of treatment effect and value.

## Data Availability

All data produced in the present study are available upon reasonable request to the authors

